# Immunogenicity and vaccine-serotype carriage prevalence after full or fractional doses of Pneumococcal Conjugate Vaccines in Kenyan infants: an individually randomised, controlled, non-inferiority trial

**DOI:** 10.1101/2024.01.24.24301730

**Authors:** Katherine E. Gallagher, Ruth Lucinde, Christian Bottomley, Mary Kaniu, Badaud Suaad, Mary Mutahi, Laura Mwalekwa, Sarah Ragab, Louise Twi-Yeboah, James A. Berkley, Mainga Hamaluba, Angela Karani, Jimmy Shangala, Mark Otiende, Elizabeth Gardiner, Daisy Mugo, Peter G. Smith, Collins Tabu, Fred Were, David Goldblatt, J. Anthony G. Scott FMedSci

**Affiliations:** Faculty of Epidemiology and Population Health, London School of Hygiene and Tropical Medicine, UK; KEMRI-Wellcome Trust Research Programme, Kilifi, Kenya; Department of Paediatrics, Coast General Teaching & Referral Hospital, Mombasa, Kenya; Great Ormond Street Institute of Child Health, University College London, UK; Centre for Tropical Medicine & Global Health, University of Oxford, UK; Immunization, UNICEF, Nairobi; School of Medicine, University of Nairobi

**Author notes:** Corresponding author: Katherine Gallagher, KEMRI Wellcome Trust Research Programme, PO Box 230, 80108, Kilifi, Kenya.

**Keywords:** randomised controlled trial, non-inferiority, fractional dosing/ dose response, pneumococcal vaccines

## Abstract

**Background:** Pneumococcal conjugate vaccines (PCVs) are the most expensive component of the routine immunisation schedule in Gavi-supported countries. As countries transition out of Gavi support, PCV programmes are at risk. We assessed whether immunogenicity was non-inferior after fractional doses of PCV10 (GlaxoSmithKline plc.) or PCV13 (Pfizer Inc.), when compared to full doses, and analysed vaccine serotype (VT) carriage prevalence.

**Methods:** 2100 healthy infants were enrolled and randomised into seven equal-sized trial arms. Doses were delivered in the 2p+1 schedule (6, 14 weeks and 9-12 months) in six trial arms: A) Full dose PCV13, B) 40%-PCV13, C) 20%-PCV13, D) Full dose PCV10, E) 40%-PCV10, F) 20%-PCV10. Participants in the seventh trial arm received full dose PCV10 at 6, 10 and 14 weeks. Immunogenicity was assessed 4-weeks post-prime and 4-weeks post-boost. Carriage was assessed at 9 and 18 months of age.

**Results:** In the per-protocol analysis, 40%-PCV13 met the non-inferiority criteria for 12/13 serotypes post-prime and 13/13 serotypes post-boost. 20%-PCV13 met the criteria for 9/13 serotypes post-prime and 10/13 serotypes post-boost. 40%-PCV10 met the criteria for 8/10 serotypes post-prime and 6/10 serotypes post-boost. 20%-PCV10 met the criteria for 7/10 serotypes post-prime and 1/10 serotype post-boost.

**Conclusions:** A 3-dose schedule of 40%-PCV13 met the non-inferiority criteria at both timepoints and could be implemented by using 4-dose UNICEF vials as 10-dose vials. A 3-dose schedule of 40%-PCV13 would cost UNICEF 3.30 USD and represents the most affordable, effective PCV schedule option currently available for countries transitioning out of Gavi-support. ClinicalTrials.gov ID: NCT03489018.

## Introduction

Pneumococcal conjugate vaccines (PCVs) have proven to be highly effective in reducing vaccine-type pneumococcal disease^1–3^. Since 2010, Gavi, the Vaccine Alliance, has funded PCV introduction in 47 ‘Gavi-eligible’ low and lower-middle-income countries (LMICs). However, even at a subsidised cost of US$2.00-3.30 per dose, the PCV programme is the most expensive component of the routine immunisation schedule in many LMICs^4^. The World Health Organization (WHO) recommends a three dose schedule: in a primary series at 6-8, 10-12 and 14-20 weeks of age (the ‘3p+0’ schedule), or two primary doses at 6-8 weeks and 14-16 weeks of age with a booster at least 6 months after the 2^nd^ dose (the ‘2p+1’ schedule)^5^. At US$6.00-9.90 per child, the sustainability of the PCV programme is of concern in countries that are transitioning out of Gavi support and are taking on the full cost of procuring the vaccine. Furthermore, for middle-income countries ineligible for Gavi support that have not yet introduced PCV, a reduction in the cost of PCV may enable vaccine introduction where it is currently unaffordable.

One approach to reducing the cost of PCV programmes is to use a fractional dose at each vaccination. Fractional doses of antigen have been shown to induce non-inferior immune responses to full doses in trials of vaccines against *Haemophilus influenzae* type b (Hib)^6–10^, *Neisseria meningitidis*^11^, Yellow Fever^12–15^ and Polio^16–20^. A systematic review identified only one published trial of a PCV at a low dose^21^. An early trial of a pentavalent PCV documented serotype-specific immune responses that reached the threshold of protection (≥0.35 mcg/ml; established following later efficacy trials) after a dose of just 0.5 mcg of antigen, without an adjuvant. This dose equates to 23% of the current dose in Prevnar13, PCV13 (Pfizer Inc.), and 50% of the dose in Synflorix, PCV10 (GlaxoSmithKline, GSK, Plc.; Table 1)^22^, though the two vaccines use different carrier proteins and conjugation methods^23,24^.

**Table 1.** The available vaccine formulations and proposed fractional (20% and 40%) dose of PCV10 (GSK) and PCV13 (Pfizer)

| Serotype-specific<br>Saccharide<br>dose (µg) | 1 | 3 | 4 | 5 | 6A | 6B | 7F | 9V | 14 | 18C | 19A | 19F | 23F |
| --- | --- | --- | --- | --- | --- | --- | --- | --- | --- | --- | --- | --- | --- |
| **PCV13<sup>1</sup>** | 2.2 | 2.2 | 2.2 | 2.2 | 2.2 | 4.4 | 2.2 | 2.2 | 2.2 | 2.2 | 2.2 | 2.2 | 2.2 |
| **40%-PCV13** | 0.88 | 0.88 | 0.88 | 0.88 | 0.88 | 1.76 | 0.88 | 0.88 | 0.88 | 0.88 | 0.88 | 0.88 | 0.88 |
| **20%-PCV13** | 0.44 | 0.44 | 0.44 | 0.44 | 0.44 | 0.88 | 0.44 | 0.44 | 0.44 | 0.44 | 0.44 | 0.44 | 0.44 |
| **PCV10<sup>2</sup>** | 1.0 |  | 3.0 | 1.0 |  | 1.0 | 1.0 | 1.0 | 1.0 | 3.0 |  | 3.0 | 1.0 |
| **40%-PCV10** | 0.40 |  | 1.2 | 0.40 |  | 0.40 | 0.40 | 0.40 | 0.40 | 1.20 |  | 1.20 | 0.40 |
| **20%-PCV10** | 0.2 |  | 0.6 | 0.2 |  | 0.2 | 0.2 | 0.2 | 0.2 | 0.6 |  | 0.6 | 0.2 |

<sup>1</sup> The saccharide in PCV13 is conjugated to CRM197 carrier protein
<sup>2</sup> The saccharide in PCV10 is conjugated to Non-Typeable H. influenzae protein D, or tetanus
toxoid (ST18C), or Diphtheria Toxin (ST19F).

We aimed to assess whether fractional doses (20% or 40%) of PCV10 or PCV13, administered in a 2p+1 schedule, induce non-inferior serotype-specific immunogenicity to the effects achieved with full doses. We also aimed to also assess the effect of the fractional dose schedule on VT carriage.

## Methods

### The study population

The trial enrolled children at 9 health facilities in Kilifi and Mombasa Counties, Kenya. PCV10 (GSK) had been introduced in the national EPI programme in Kenya in 2011 in a 3p+0 schedule. In 2017, coverage with the third dose of PCV10 among children aged 12-23 months was 89%^25^. There was evidence of substantial indirect protection against carriage and disease for unvaccinated members of the population^2^. In 2016, there were 3.2 cases of vaccine-type invasive pneumococcal disease (IPD) per 100,000 person years in children aged <5 years^26^; surveillance for IPD continued throughout the trial period.

### Trial design

Anticipating a potential change in the schedule, we designed the study to deliver PCV in a 2p+1 schedule and delivered the primary doses at 6 and 14 weeks of age. We co-administered the PCV booster dose with MCV1 at 9 months of age. MCV1 coverage was approximately 78% in 2017^27^. At 6-8 weeks of age, 2100 infants were randomised equally into seven trial arms and followed until 18 months of age. Six trial arms delivered PCV in a 2p+1 schedule: Arm A) Full dose PCV13 B) 40% fractional dose PCV13 C) 20% fractional dose PCV13 D) Full dose PCV10 E) 40% fractional dose PCV10 F) 20% fractional dose PCV10. The seventh trial arm, Arm G, delivered PCV10 in a 3p+0 schedule to bridge the findings in the experimental arms to the existing dose and schedule in Kenya **(Figure 1).**

**Figure 1.**
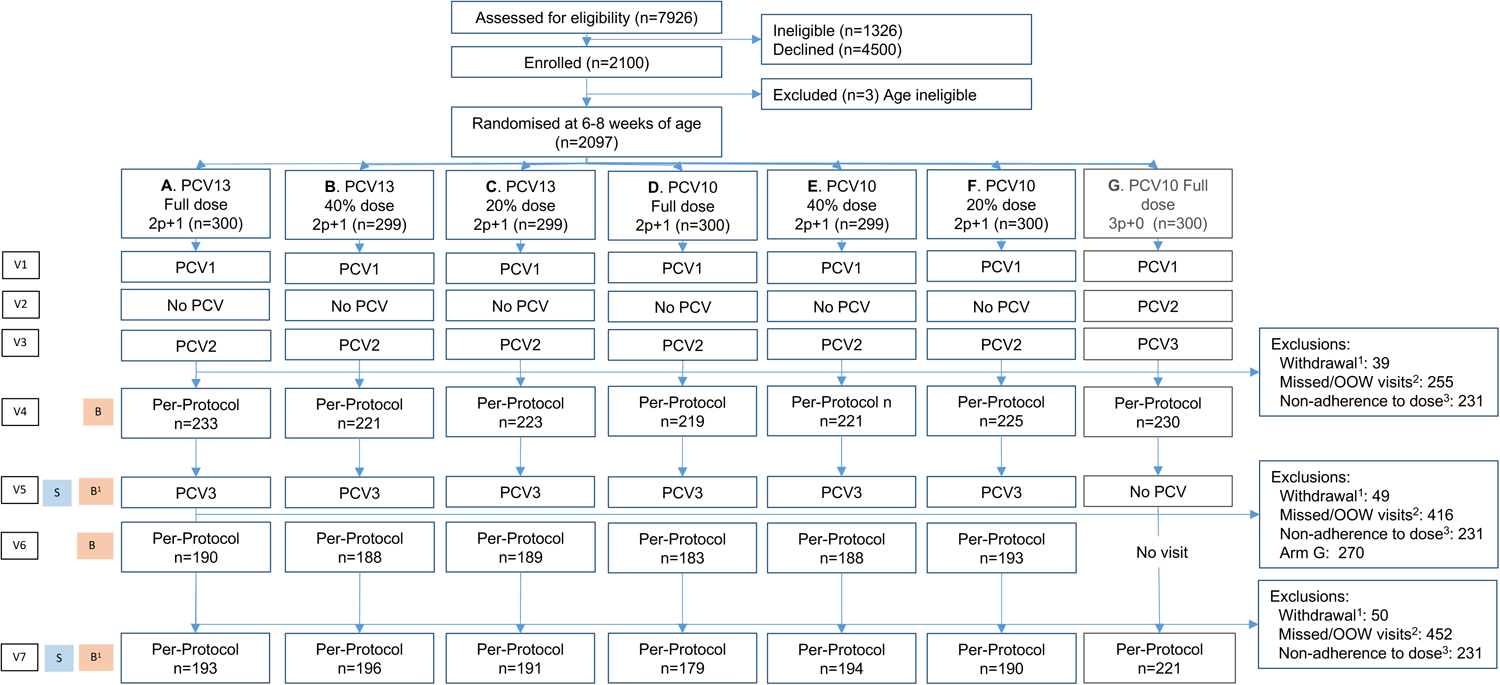
The clinical trial flow. ^1^Withdrawals include parental withdrawals, investigator-initiated withdrawals, deaths and 3 participants who were identified as ineligible for the study after randomization; ^2^Missed or out of window visits refer to missed or out of window vaccination or sampling visits, and missed samples; ^3^Non-adherence to dose includes participants who received full doses outside of the study activities (predominantly during the pause in research activities during the COVID-19 pandemic), and 13 randomization and vaccination errors. Abbreviations: B: Blood sample (Half of the participants in Arms A-F were randomly assigned to blood draws at V5 and V7); OOW: out-of-window (visit 4 and visit 6 had to occur 28 days after the last PCV dose, all visit windows were +/− 7days apart from visit 5 which could occur between D214 and D312, and visit 7 (+/− 14 days); PCV: Pneumococcal Conjugate Vaccine (doses 1-3); S: Nasopharyngeal swab; V1: visit 1 at Day 0; V2: visit 2 at Day 28 +/− 7 days; V3: visit 3 at Day 56 +/−7 days; V4: visit 4 at 28 days post-V3 +/− 7 days; V5: visit 5 at Day 228 +3months/-2weeks; V6: visit 6 at 28 days post-V5 +/− 7days; V7: visit 7 at Day 502 +/− 14 days.

### Study procedures

Communities were engaged with information about the trial. Community Health Volunteers identified households with newborns and brought the carers to the health facility, where staff provided information about the study. We recruited any healthy infant (i.e. no acute febrile illness on the day of enrolment) aged 6-8 weeks, who was eligible for vaccination in the routine immunisation programme but had not yet received their first dose of PCV. Infants more than 8 weeks of age, or those with acute illness at the time of enrolment were excluded from the trial. Less than 5% of mothers were HIV infected **(Suppl. Table 2B)** and infants were enrolled irrespective of HIV status.

Each infant was randomly allocated to one of the seven trial arms with equal probability using sequentially numbered, sealed envelopes. Computer-generated randomisation codes were prepared in advance by an independent statistician using block sizes of 14. Parents of participants in arms A-F were blinded to the dose allocated. Other than the team administering vaccine, all other study personnel were blinded to allocation of participants until the end of the study. Blinding of arm G, full dose PCV10 in a 3p+0 schedule, was not possible. A research nurse prepared 0.5ml as a full dose, 0.2ml as a 40% dose or 0.1ml as a 20% dose and administered the vaccine, in a masked syringe, intramuscularly in the right anterolateral thigh muscle. Participants received other immunisations according to the routine schedule.

Participants were invited back to the clinic at 28 days and 56 days post-enrolment, the primary series of PCV was administered according to the child’s allocated schedule. Four weeks after visit 3, at approximately 18 weeks of age, a blood sample was collected from all infants. At approximately 9 months of age (visit 5) blood was sampled from half of the participants in each of arms A-F, as allocated during randomisation, and a single nasopharyngeal swab was collected from all participants. Infants in arms A-F also received their third, booster, dose of PCV. Four weeks after the booster dose (visit 6), a post-boost blood sample was collected from all participants in arms A-F.

Finally, at approximately 18 months of age (visit 7), a nasopharyngeal swab was taken from all and a blood sample was taken from half of the participants in each of the trial arms A-F, as allocated during randomisation. Between scheduled visits participants attended the clinic at any time to receive medical care for acute illnesses. A blinded member of the study team assessed each child 7 days after each PCV dose for injection site abscesses.

Adverse events (AEs) and serious adverse events (SAEs) were defined in accordance with the International Conference on Harmonization (ICH) Guidelines for Good Clinical Practice. AEs were treated by the unblinded study nurses stationed at each facility. All SAEs were treated by a blinded study clinician at hospital.

### Laboratory methods

A maximum of 2ml of whole blood was collected via venepuncture and transported to the KEMRI-Wellcome Trust Research Programme (KWTRP) laboratory at 2-8 degrees; serum was separated within 48 hours. One aliquot from each visit was shipped to the UCL WHO Reference Laboratory for Pneumococcal Serology based at the Great Ormond Street Institute of Child Health. At UCL, sera were assayed for IgG to vaccine-type capsular polysaccharides using the WHO reference ELISA (http://www.vaccine.uab.edu/ELISA%20Protocol.pdf) and, on a subset (n=50, 1-month post-boost), for functional antibody using the multiplexed Opsonophagocytic Assay (MOPA; http://www.vaccine.uab.edu/UAB-MOPA.pdf)^28^. Samples were analysed for IgG to VTs, except for samples from the routine immunisation arm (PCV10 3p+0) which were assayed for 7 VTs.

Standard methods for collecting and processing nasopharyngeal swabs for culture were used to ensure that results are comparable with previous studies^29,30^. A single nasopharyngeal swab was taken at 9 and 18 months of age and transported in 1 ml skim milk-tryptone-glucose-glycerin (STGG) transport medium to the KWTRP CGMR-Coast laboratories. A primary culture was prepared on blood agar with gentamicin, one colony on the plate was selected at random for serotyping by latex agglutination and confirmatory Quellung reaction. Polymerase Chain Reaction (PCR) was performed for quality control purposes on 10% of the samples, and, as a confirmatory test for samples that had ambiguous or negative Quellung tests. VT carriage was defined if a vaccine serotype was isolated by latex agglutination and confirmatory Quellung reaction^30^.

### Statistical analyses

The assessment of non-inferiority was based on the lower limit of a 2-sided 90% confidence interval, which is equivalent to the limit of a one-sided 95% confidence interval. At 4-weeks post-boost, non-inferiority was declared if the lower limit of the 90% CI for the ratio of geometric mean concentration (GMC) of IgG (fractional/full dose arms) was more than 0.5 (i.e. the less than 2-fold reduction criterion). At 4 weeks after the primary series (18 weeks of age), the proportion of ‘responders’, was defined as those with serotype-specific IgG antibody concentration ≥0.35 mcg/ml^31^, and non-inferiority was declared if the lower limit of the 90% CI around the difference in the proportion of ‘responders’ (fractional dose group – full dose group) was >-10%^32^. *A priori*, a vaccine dose was defined as non-inferior if it met the non-inferiority criteria for at least 8 of the 10 vaccine types in the PCV10 arms and at least 10 of the 13 vaccine types in the PCV13 arms.

Analyses were restricted to the per-protocol population i.e. randomized participants in arms A-G who received three doses of their allocated dose in the allocated schedule and had at least one blood sample within window (for immunogenicity analyses) or at least one carriage sample within window (for carriage analyses). No interim analyses were conducted.

The required sample size for the trial was calculated to ensure sufficient power for both the 18-week and 10-month assessments of non-inferiority^33^. We assumed serotype-specific proportions of children reaching the correlate of protection (IgG <u>≥</u>0.35 mcg/ml, on average 87%) at 18 weeks of age^34–38^. To declare non-inferiority at the 18-week assessment with 90% power, we estimated that we would need to enrol 300 infants per arm, accounting for serotype-specific response rates and 5% loss to follow up. This sample size conferred >99% power to declare non-inferiority in the analyses post-boost, assuming similar GMCs to those elicited after a 2p+1 schedule in South African children^38^.

Ethics approval was obtained from Kenyan Medical Research Institute Scientific & Ethics Review Unit (SERU) and the London School of Hygiene & Tropical Medicine (LSHTM) Ethics Committee. Written informed consent was obtained from at least one caregiver of all infants enrolled in the study. An independent data and safety monitoring committee provided trial oversight.

## Results

Between March 2019 and November 2021, 2100 infants and their parents were enrolled into the trial; 673 (32%) participants were enrolled between March 2019 - March 2020, trial activities were then paused for 7 months due to the COVID-19 pandemic; 1427 infants were enrolled between October 2020 and November 2021 **(Figure 1)**. Due to the disruption of follow-up in 2020, 1572 participants out of 2100 (75%) were included in the per-protocol analysis at 18 weeks of age (**Suppl. Table 1A**). For the post-boost immunogenicity analysis at approximately 10 months of age, 1131 participants of the 1797 (63%) who were allocated to 2p+1 arms were included in the per-protocol analysis (**Suppl. Table 1B**). For the carriage prevalence analyses, 1439 participants of the 2100 enrolled (69%) were included in the per-protocol analysis at 9 months of age and 1364 (65%) at 18 months of age (**Suppl. Table 1C & 1D**). The characteristics of participants in the per-protocol analysis were balanced across the arms with respect to sex, HIV exposure (maternal HIV status), infant weight at enrolment, breastfeeding at 10 months of age and the timing of their boost dose **(Suppl. Table 2A, 2B, 2C, 2D**).

### Immunogenicity at 18 weeks, post-primary series

Compared to 2 full doses of PCV13, the noninferiority criterion was met for 12 of the 13 serotypes in the 40% PCV13 recipients, but for only 9 of the 13 serotypes in the 20% PCV13 recipients. Compared to 2 full doses of PCV10, the non-inferiority criterion was met for 8 of the 10 serotypes in the 40% PCV10 recipients, but for only 7 of the 10 STs in the 20% PCV10 recipients (**Figure 2**). A primary series of 2 PCV10 doses was non-inferior to a primary series of 3 PCV10 doses for 6 of the 7 serotypes assayed, except 23F (**Suppl. Tables 3A, 3B**).

**Figure 2.**
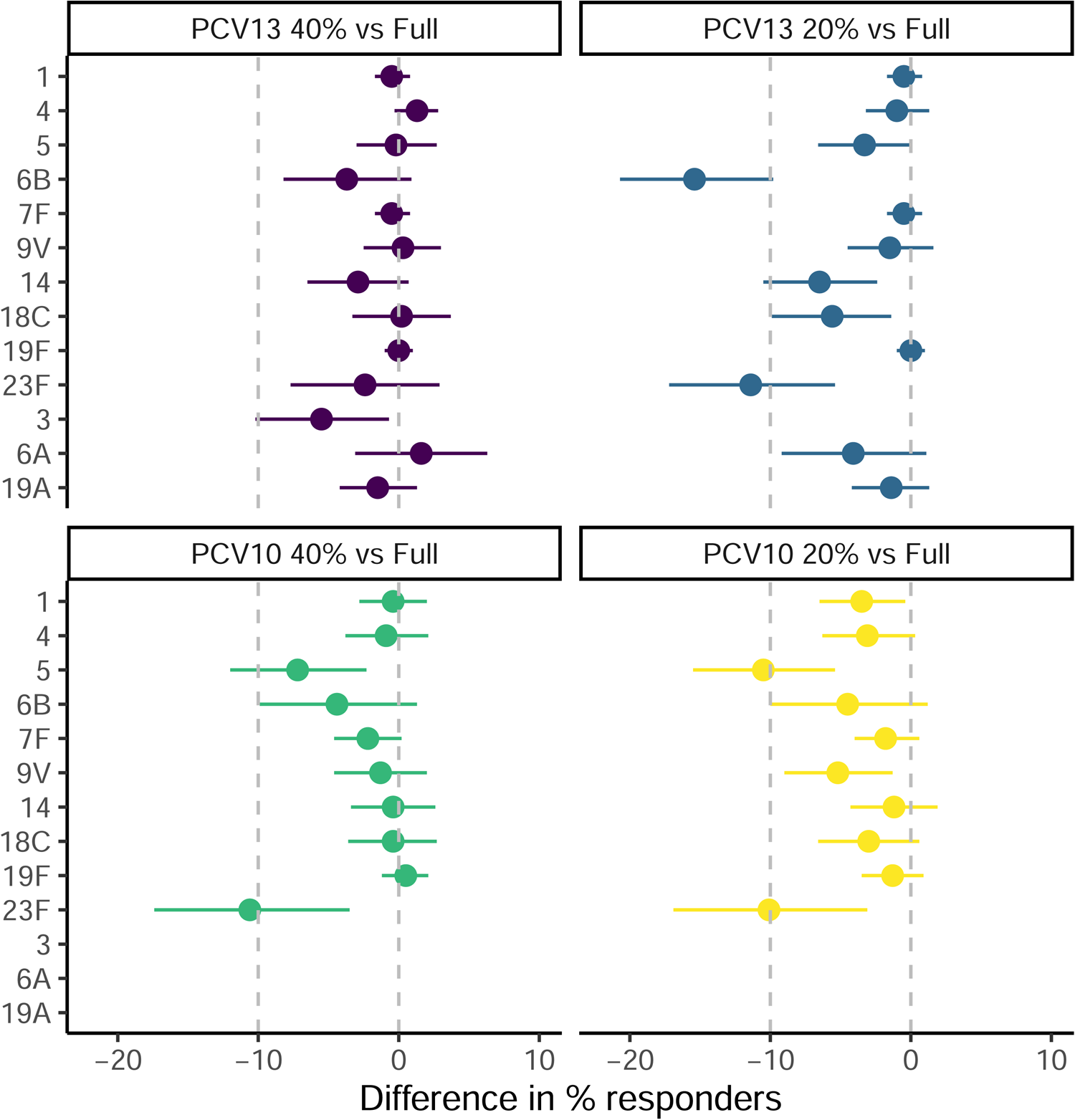
The difference in the proportion of responders (full dose-fractional dose) at 4-weeks post primary series (18 weeks of age) Notes: The difference in the proportion of responders is missing for the comparison of full dose PCV13 and 20% PCV13, it is larger than the x-axis scale (−30.4, 90%CI −36.3, −24.0; suppl. Table 3B).

### Immunogenicity at 10 months, post-boost

In the 2p+1 arms, compared to full dose PCV13 recipients, the post-boost noninferiority criterion was met for 13 of the 13 serotypes in the 40% PCV13 recipients, and for 10 of the 13 serotypes in the 20% PCV13 recipients. In the 2p+1 arms, compared to full dose PCV10 recipients, the post-boost non-inferiority criterion was only met for 6 of the 10 serotypes in the 40% PCV10 recipients and only 1 of the 10 STs in the 20% PCV10 recipients **(Figure 3; Suppl. Tables 4A, 4B**). In the sub-set of participants who had a sample assayed for opsonophagocytic function, the proportion of samples with geometric mean OPA titres of >8 was high across all arms and serotypes **(suppl. Table 4C, 4D, 4E**).

**Figure 3.**
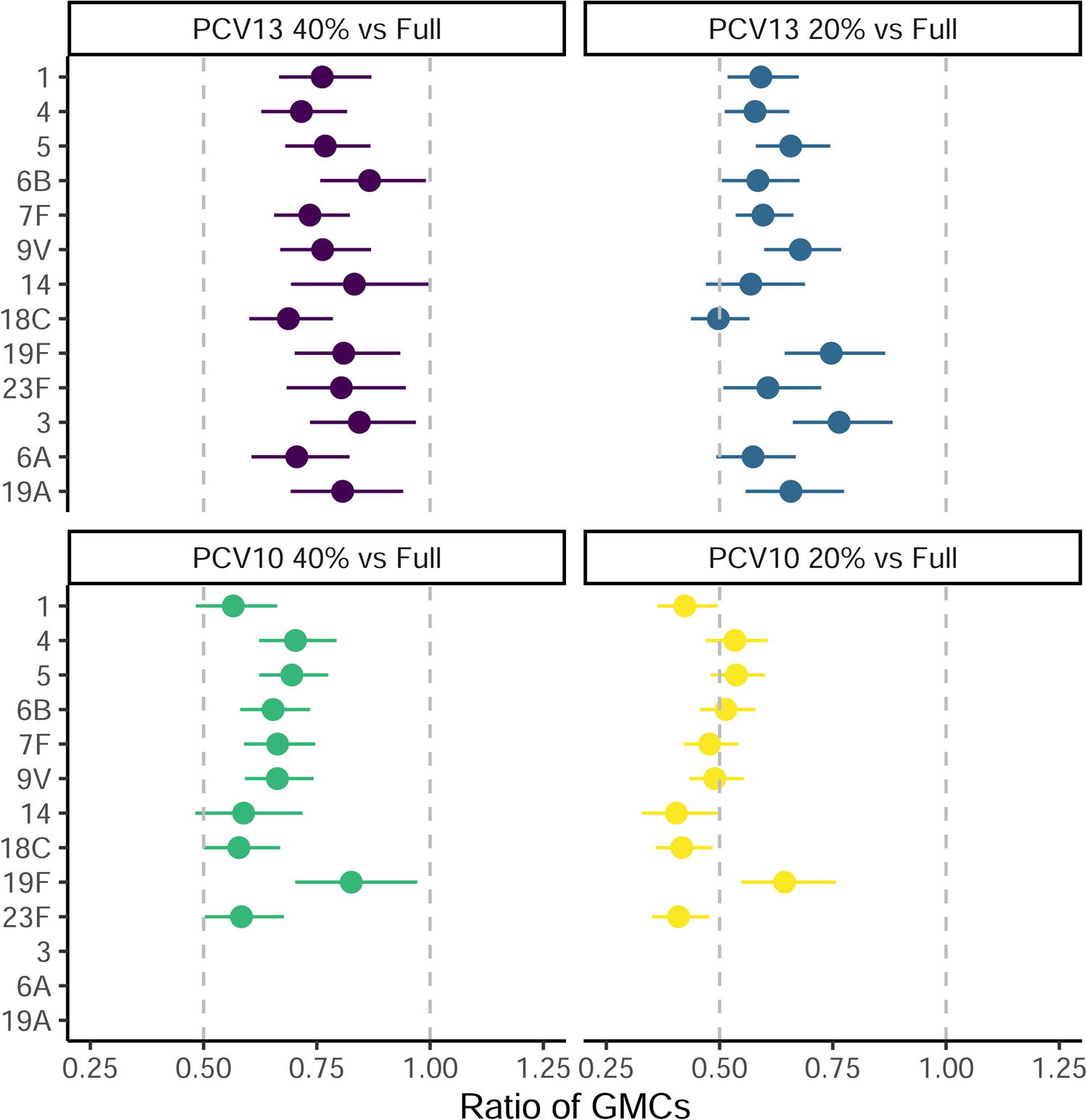
The ratio of GMCs post-boost (full dose: fractional dose; approximately 10 months of age)

### Carriage prevalence

There were no statistically significant differences (p<0.05) in vaccine type carriage prevalence by dose or by number of doses in the primary series (2 vs 3) at 9 months of age (**Table 2; suppl. Table 5A, 5B**). At 18 months of age, there were no statistically significant differences in vaccine type carriage prevalence by dose of PCV13; however, the full dose 2p+1 PCV10 recipients had significantly lower PCV10 VT carriage prevalence than the 20% PCV10 recipients (p=0.008). Additionally, the full dose PCV10 2p+1 arm had significantly lower PCV10 type carriage than the full dose PCV10 3p+0 arm (p=0.016, **Table 2; Suppl. Table 5C, 5D**).

**Table 2.**
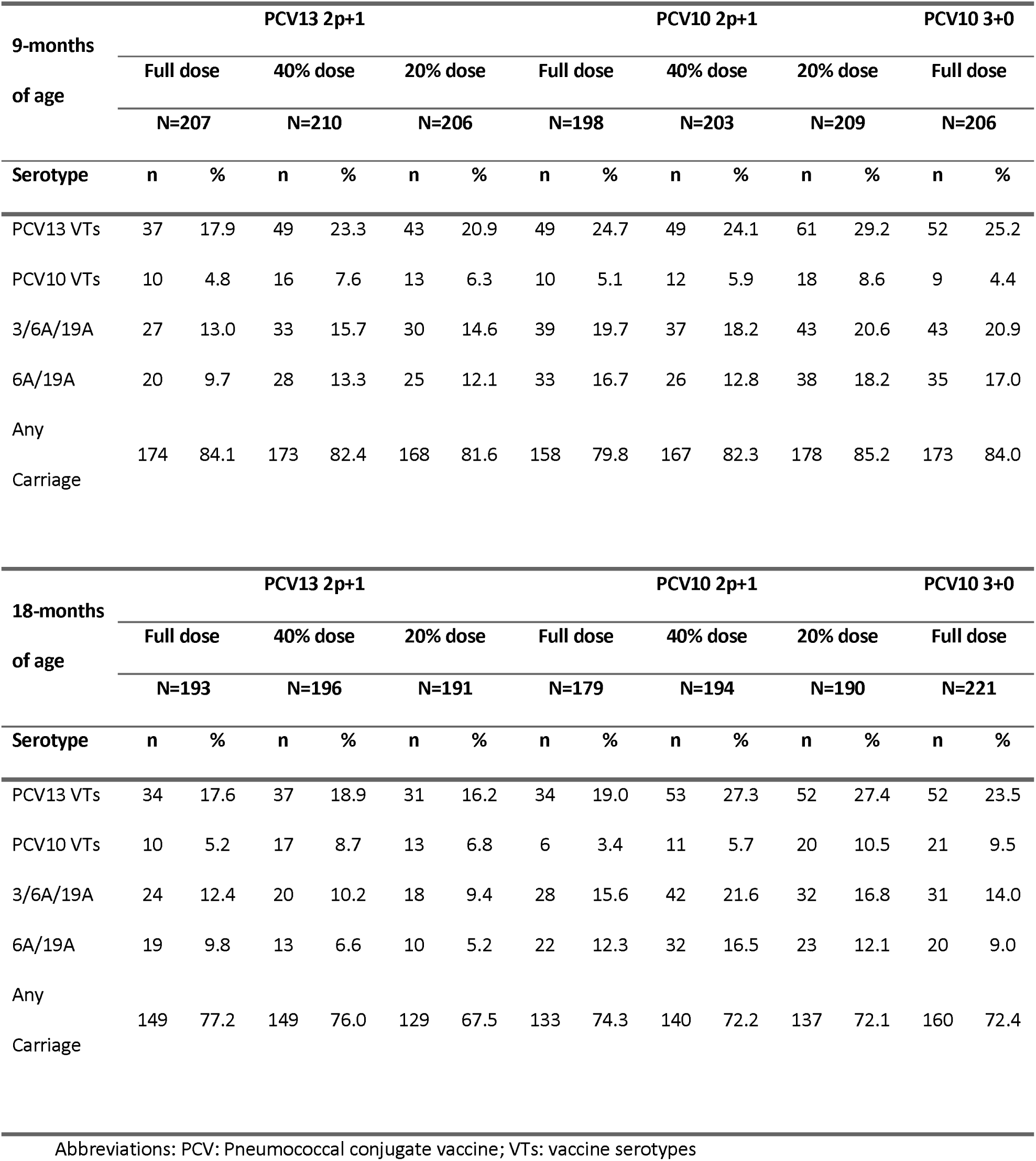
Carriage prevalence at 9 & 18 months of age.

### Safety

No injection site abscesses were recorded. A total of 61 cases of non-severe pneumonia and 65 SAEs were recorded, these were distributed across the arms evenly **(Suppl. Table 6**).

## Discussion

A 2p+1 schedule of 40% doses of PCV13 elicited non-inferior IgG responses after both the primary series and the booster dose, when compared to a full dose 2p+1 schedule. Smaller, 20%, doses of PCV13 elicited inferior immunogenicity after the primary series but met the non-inferiority criteria after the boost, when compared to a full dose schedule. There were no significant differences in VT carriage prevalence across the PCV13 arms at 9 nor 18 months of age. The per-protocol populations for the non-inferiority analyses were smaller than planned, which is likely to have reduced the precision with which we could estimate the ratios of the proportion of responders and of GMCs. The 20% PCV13 arm narrowly missed the non-inferiority criteria for ST14 and ST18, post-primary series, and may have met the criteria with a larger sample size.

Fractional dose schedules of PCV10 failed to meet non-inferiority criteria for immunogenicity and demonstrated statistically significantly higher VT carriage prevalence at 18 months of age when compared to the full dose arm. These results align with a dose-response relationship across products: full dose PCV13, full dose PCV10 and 40% PCV13 contain at least 0.88ug of saccharide, whereas 20% PCV13, 20% PCV10 and 20%PCV10 contain less than 0.88ug (Table 1). A schedule of 2 full primary doses of PCV10 elicited noninferior immunogenicity to 3 full primary doses of PCV10 among 6 of the 7 ST-specific responses that were assayed (all except ST23F) at 18 weeks of age. The 2p+1 arm had significantly lower VT carriage prevalence than the 3p+0 arm at 18 months of age.

The Government of Kenya has announced it aims to fully self-finance its routine immunisation programme by 2030. In 2022, the country switched from the PCV10 produced by GlaxoSmithKline (US$3.05 per dose), to the more economical alternative produced by Serum Institute of India (SII; US$2.00 per dose). A 3-dose schedule of 40% PCV13 (US$ 1.1 per dose) represents the most affordable option on the market currently and could reduce the annual cost of purchasing PCV for an annual birth cohort of 1.5 million children, from 9 to 5 million USD (at the current costs, assuming no vaccine wastage). Furthermore, 4-dose vials of PCV13 are available, and contain a preservative, which reduces the risk of contamination and enables multi-dose vials to be used for up to 28 days following first puncture. It would be feasible, therefore, to implement a 40% dose policy immediately by re-classifying the present 4-dose vials of PCV13 as a 10-dose vials of 40%-doses (0.2ml).

The SII PCV10 contains the same amount of saccharide as PCV13, but due to differences in the manufacturing processes we cannot assume our findings are generalisable to the SII PCV10. At the time of trial design, only the GSK PCV10 and the PCV13 were available for introduction into the routine immunisation schedules of LMICs and we could not study how the new, higher valency vaccines on the market perform at fractional doses.

At both immunogenicity timepoints, we used non-inferiority criteria that conform with those used in vaccine licensure studies and the threshold of protection that is used for licensure. There is some evidence that the threshold of protection against IPD varies by serotype^39^, and that correlates of protection against carriage are substantially higher than those against invasive disease^40^. It is unclear whether the lower, albeit non-inferior, immunogenicity of 40% PCV13, would influence protection against carriage acquisition or against pneumococcal disease. We did not detect increased acquisition during our study; however, our trial population was under substantial indirect protection conferred by the high coverage of PCV in the routine immunisation system.

None of the children enrolled on the study acquired HIV, although a small number were exposed to the virus. The study was not designed to determine whether HIV-infected infants would mount a protective response following a schedule of 40% PCV13. In Kenya, HIV prevalence in women with at least 1 child was 6.1% in 2014 and it is estimated that 0.9% of infants have HIV^41^.

In conclusion, 40% doses of PCV13 in a 2p+1 schedule generated non-inferior immune responses and no difference in vaccine type carriage prevalence when compared to a full dose schedule. Although the long-term impact of switching to a 40% PCV13 schedule on carriage transmission, and the generalisability of findings to HIV+ populations, are both unclear, a 3-dose schedule using 40% PCV13 is currently the most economically effective option for PCV programmes in Gavi-transitioning LMICs and middle-income countries unsupported by Gavi.

## Data Availability

All data produced in the present study are available upon reasonable request to the authors

## Acknowledgements

We would like to thank: the families of all the participants for contributing a substantial amount of time to attending the trial visits; the KEMRI-CGMR-C Community Liaison Group for supporting extensive community engagement activities throughout 2018-2021; the Ministry of Health staff at the County and sub-county levels for their leadership and advice; the hospital/ health facility management committees and the health facility staff who supported trial activities despite many set-backs and delays especially during the COVID-19 pandemic. We would also like to thank the dedicated team of nurses, fieldworkers, clinicians, data managers and laboratory scientists who worked on the trial. We thank the data safety and monitoring board for consistently reviewing progress reports, accruing safety data and the primary endpoint results.

This paper is published with the permission of the Director of KEMRI.

## Author contributions

Katherine E. Gallagher: funding acquisition, study design, project administration, supervision, writing - original draft

Christian Bottomley: formal analysis, writing – review & editing

Ruth Lucinde, Mary Kaniu, Badaud Suaad, Laura Mwalekwa, Elizabeth Gardiner, Daisy Mugo, Mark Otiende, Sarah Ragab, Louise Twi-Yeboah, Angela Karani, Jimmy Shangala: Investigation, writing – review & editing

Mary Mutahi: data curation, writing – review & editing

James A. Berkley, Mainga Hamaluba: writing – review & editing, site oversight

Peter G. Smith, Collins Tabu, Fred Were: study design, writing – review & editing.

David Goldblatt: resources, supervision, writing – review & editing

J. Anthony G. Scott: conceptualisation, funding acquisition, study design, supervision, writing – review & editing

## Trial Registration

ClinicalTrials.gov ID: NCT03489018; Pan African Clinical Trial Registry ID: PACTR202104717648755.

Full trial protocol can be accessed as supplementary information to this manuscript

## Funding acknowledgement

The Bill & Melinda Gates Foundation, and The UK National Institute of Health Research (NIHR) Global Health Research Unit on Mucosal Pathogens (MPRU).

