## Supplementary files for "Immunogenicity and vaccine-serotype carriage prevalence after full or fractional doses of Pneumococcal Conjugate Vaccines in Kenyan infants: an individually randomised, controlled, non-inferiority trial"

**Supplementary Tables**

**Table S1A: Exclusions from the post-primary series (18-week of age) per-protocol population (immunogenicity)**

| | PCV13 2p+1 | | | PCV10 2p+1 | | | PCV10 3+0 | |
|---|---|---|---|---|---|---|---|---|
| | **Full dose** | **40% dose** | **20% dose** | **Full dose** | **40% dose** | **20% dose** | **Full dose** | **All** |
| Per-protocol cohort | 233 | 221 | 223 | 219 | 221 | 225 | 230 | 1,572 |
| Ineligible[1] | 1 | 0 | 0 | 0 | 2 | 0 | 0 | 3 |
| Randomisation error[2] | 0 | 0 | 2 | 3 | 3 | 2 | 3 | 13 |
| Received PCV dose outside study[3] | 31 | 39 | 26 | 29 | 38 | 33 | 22 | 218 |
| 2nd PCV dose missed or not in window | 12 | 14 | 16 | 21 | 15 | 17 | 12 | 107 |
| Withdrew from study | 6 | 6 | 6 | 4 | 5 | 2 | 5 | 34 |
| Died | 1 | 0 | 0 | 1 | 0 | 0 | 0 | 2 |
| 18-week visit missed or not in window | 12 | 13 | 20 | 11 | 11 | 15 | 21 | 103 |
| Sample stored >48 hrs after collection[4] | 0 | 0 | 1 | 0 | 0 | 0 | 0 | 1 |
| Sample missing[5] | 4 | 6 | 5 | 12 | 4 | 6 | 7 | 44 |
| Total | 300 | 299 | 299 | 300 | 299 | 300 | 300 | 2,097 |

**Table S1B: Exclusions from the post-boost (approx. 10-month of age) per-protocol population (immunogenicity)**

| | PCV13 2p+1 | | | PCV10 2p+1 | | | PCV10 3+0 | |
|---|---|---|---|---|---|---|---|---|
| | **Full dose** | **40% dose** | **20% dose** | **Full dose** | **40% dose** | **20% dose** | **Full dose** | **All** |
| Per-protocol cohort | 190 | 188 | 189 | 183 | 188 | 193 | 0 | 1,131 |
| Ineligible[1] | 1 | 0 | 0 | 0 | 2 | 0 | 0 | 3 |
| Randomisation error[2] | 0 | 0 | 2 | 3 | 3 | 2 | 3 | 13 |
| Received PCV dose outside study[3] | 31 | 39 | 26 | 29 | 38 | 33 | 22 | 218 |
| 2nd PCV dose missed or not in window | 12 | 14 | 16 | 21 | 15 | 17 | 12 | 107 |
| 3rd PCV dose missed or not in window[6] | 39 | 28 | 39 | 44 | 33 | 34 | 258 | 475 |
| Withdrew from study | 8 | 7 | 8 | 4 | 5 | 3 | 5 | 40 |
| Died | 1 | 1 | 1 | 1 | 0 | 2 | 0 | 6 |
| Post-boost visit missed or not in window | 14 | 13 | 12 | 13 | 11 | 12 | 0 | 75 |
| Sample stored >48 hrs after collection[4] | 0 | 0 | 0 | 1 | 0 | 0 | 0 | 1 |
| Sample missing[5] | 4 | 9 | 6 | 1 | 4 | 4 | 0 | 28 |
| Total | 300 | 299 | 299 | 300 | 299 | 300 | 300 | 2,097 |

**Table S1C: Exclusions from the 9-month of age per-protocol population (carriage)**

| | PCV13 2p+1 | | | PCV10 2p+1 | | | PCV10 3+0 | |
|---|---|---|---|---|---|---|---|---|
| | **Full dose** | **40% dose** | **20% dose** | **Full dose** | **40% dose** | **20% dose** | **Full dose** | **All** |
| Per-protocol cohort | 207 | 210 | 206 | 198 | 203 | 209 | 206 | 1,439 |
| Ineligible[1] | 1 | 0 | 0 | 0 | 2 | 0 | 0 | 3 |
| Randomisation error[2] | 0 | 0 | 2 | 3 | 3 | 2 | 3 | 13 |
| Received PCV dose outside study[3] | 31 | 39 | 26 | 29 | 38 | 33 | 22 | 218 |
| 2nd PCV dose missed or not in window | 12 | 14 | 16 | 21 | 15 | 17 | 10 | 105 |
| 3rd PCV dose missed or not in window[6] | 39 | 28 | 39 | 44 | 33 | 34 | 6 | 223 |
| Withdrew from study | 8 | 7 | 8 | 4 | 5 | 3 | 3 | 38 |
| Died | 1 | 1 | 1 | 1 | 0 | 2 | 0 | 6 |
| NPS sample missing | 0 | 0 | 0 | 0 | 0 | 0 | 13 | 13 |
| NPS collection out of window | 1 | 0 | 1 | 0 | 0 | 0 | 37 | 39 |
| Total | 300 | 299 | 299 | 300 | 299 | 300 | 300 | 2,097 |

**Table S1D. Exclusions from the 18-month of age per-protocol population (carriage)**

| | PCV13 2p+1 | | | PCV10 2p+1 | | | PCV10 3+0 | |
|---|---|---|---|---|---|---|---|---|
| | Full dose | 40% dose | 20% dose | Full dose | 40% dose | 20% dose | Full dose | All |
| Per-protocol cohort | 193 | 196 | 191 | 179 | 194 | 190 | 221 | 1,364 |
| Ineligible[1] | 1 | 0 | 0 | 0 | 2 | 0 | 0 | 3 |
| Randomisation error[2] | 0 | 0 | 2 | 3 | 3 | 2 | 3 | 13 |
| Received PCV dose outside study[3] | 31 | 39 | 26 | 29 | 38 | 33 | 22 | 218 |
| 2nd PCV dose missed or not in window | 12 | 14 | 16 | 21 | 15 | 17 | 10 | 105 |
| 3rd PCV dose missed or not in window[6] | 39 | 28 | 39 | 44 | 33 | 34 | 6 | 223 |
| Withdrew from study | 8 | 7 | 8 | 4 | 5 | 3 | 6 | 41 |
| Died | 1 | 1 | 1 | 1 | 0 | 2 | 0 | 6 |
| NPS sample missing | 0 | 0 | 0 | 0 | 0 | 2 | 0 | 2 |
| NPS collection out of window | 15 | 14 | 16 | 19 | 9 | 17 | 32 | 122 |
| Total | 300 | 299 | 299 | 300 | 299 | 300 | 300 | 2,097 |

Table 1A, 1B, 1C, 1D notes:

[1] Three participants were age-ineligible at enrolment (<6 weeks) and this was only checked and determined post-randomisation. Protocol violations were submitted for the incorrect enrolment and randomisation of these participants. An additional three participants were enrolled however were ineligible with a fever on physical examination and were not randomized – these participants therefore do not feature in this table, explaining the total of 2097.

[2] Randomisation/ vaccination errors occurred when staff identified the incorrect randomisation code for the participant and the participant received a product or dose that was not allocated. Protocol violations were submitted for these deviations from SOPs.

[3] Study procedures and follow-up had to pause between 20th March 2020 and 21st September 2020 due to the COVID-19 pandemic restrictions on working. During this time, we advised participants who had not completed their primary series of vaccinations to attend their local vaccination clinic to receive a full dose of PCV, as it was unclear when we would be able to access them to complete their primary series. This led to 205 participants of the 637 enrolled before 20th March 2020 receiving a *full* second dose of PCV rather than their allocated dose. A further 13 participants of the 1463 participants enrolled after the pandemic restrictions had been lifted, received a full second dose of PCV at a different vaccination clinic rather than attending the study clinic. There were measures in place to prevent this happening including: stapling a research participation ID card to the mother-child handbook, placing a sticker stating ‘do not vaccinate’ on the PCV vaccination record page with a contact phone number, and regular telephone calls to mothers to remind them to come back to the study clinic for their vaccinations rather than go elsewhere. At each visit, a staff member would ask the mother whether their child had received any injections since they saw the study nurse and check the mother-child booklet for any indication of a vaccine dose from another vaccination clinic. If there was any uncertainty e.g., the mother reported attending another clinic and getting a vaccine but a dose was not documented in the MCH book, a staff member visited the clinic the mother reported attending and sought the clinic’s vaccination register book to confirm if a PCV dose was administered to the study participant. These 13 deviations were identified in this way either by report, documentation or follow up at neighbouring clinics.

[4] Two protocol violations were submitted for an incident whereby inadequate hand-over between lab staff led to one V4 and one V6 sample being misplaced and only stored >48 hours after sample collection.

[5] In some cases it was not possible to take a blood sample due to difficulty in locating an adequate vein, or due to parental refusal, these were submitted as protocol deviations.

[6] During the COVID-19 pandemic the window for the boost (3rd dose) visit was widened to 9-12 months of age; however, the pandemic still had a significant impact on attendance of the boost dose visit within-window: 249 of the 637 (39%) enrolled before the pandemic either missed their boost dose or attended the visit out-of-window, in-comparison to 226 of the 1463 (16%) participants enrolled after the COVID-19 restrictions had been lifted. The PCV10 3p+0 arm was excluded from the per-protocol population at 10-months of age as participants did not receive a boost dose as per the allocated schedule.

**Table S2A: Baseline characteristics of the post-primary (18-week of age) per-protocol cohort (n, %)**

| | PCV13 2p+1 | | | | | | PCV10 2p+1 | | | | | | PCV10 3+0 | |
|---|---|---|---|---|---|---|---|---|---|---|---|---|---|---|
| | Full dose | | 40% dose | | 20% dose | | Full dose | | 40% dose | | 20% dose | | Full dose | |
| | n | % | n | % | n | % | n | % | n | % | n | % | n | % |
| Season | | | | | | | | | | | | | | |
| Jan-Mar | 23 | 10 | 25 | 11 | 22 | 10 | 25 | 11 | 23 | 10 | 25 | 11 | 24 | 10 |
| Apr-Jun | 66 | 28 | 67 | 30 | 66 | 30 | 66 | 30 | 62 | 28 | 67 | 30 | 62 | 27 |
| Jul-Sep | 65 | 28 | 53 | 24 | 56 | 25 | 60 | 27 | 68 | 31 | 60 | 27 | 64 | 28 |
| Oct-Dec | 79 | 34 | 76 | 34 | 79 | 35 | 68 | 31 | 68 | 31 | 73 | 32 | 80 | 35 |
| Year | | | | | | | | | | | | | | |
| 2019 | 44 | 19 | 38 | 17 | 44 | 20 | 43 | 20 | 40 | 18 | 42 | 19 | 49 | 21 |
| 2020 | 4 | 2 | 4 | 2 | 5 | 2 | 3 | 1 | 5 | 2 | 3 | 1 | 3 | 1 |
| 2021 | 185 | 79 | 179 | 81 | 174 | 78 | 173 | 79 | 176 | 80 | 180 | 80 | 178 | 77 |
| Sex | | | | | | | | | | | | | | |
| Male | 116 | 50 | 104 | 47 | 117 | 52 | 114 | 52 | 107 | 48 | 106 | 47 | 118 | 51 |
| Female | 117 | 50 | 117 | 53 | 106 | 48 | 105 | 48 | 114 | 52 | 119 | 53 | 112 | 49 |
| Infant HIV | | | | | | | | | | | | | | |
| Positive | 0 | 0 | 0 | 0 | 0 | 0 | 0 | 0 | 0 | 0 | 0 | 0 | 0 | 0 |
| Negative | 225 | 97 | 213 | 96 | 210 | 94 | 213 | 97 | 215 | 97 | 215 | 96 | 222 | 97 |
| Unknown | 8 | 3 | 8 | 4 | 13 | 6 | 6 | 3 | 6 | 3 | 10 | 4 | 8 | 3 |

**Table S2B. Baseline characteristics of the post-primary (18-week of age) per-protocol cohort (median, inter-quartile range)**

| | PCV13 2p+1 | | | | | | PCV10 2p+1 | | | | | | PCV10 3+0 | |
|---|---|---|---|---|---|---|---|---|---|---|---|---|---|---|
| | Full dose | | 40% dose | | 20% dose | | Full dose | | 40% dose | | 20% dose | | Full dose | |
| | Med. | IQR | Med. | IQR | Med. | IQR | Med. | IQR | Med. | IQR | Med. | IQR | Med. | IQR |
| Maternal age, yrs | 25 | 21,31 | 26 | 21,31 | 26 | 21,32 | 25 | 21,30 | 27 | 21,31 | 26 | 22,31 | 27 | 22,32 |
| Infant weight, kgs | 4 | 4,5 | 4 | 4,5 | 5 | 4,5 | 4 | 4,5 | 5 | 4,5 | 4 | 4,5 | 4 | 4,5 |
| Dose 1, age in days | 43 | 42,45 | 43 | 42,46 | 44 | 42,46 | 44 | 42,46 | 43 | 42,45 | 43 | 42,45 | 44 | 42,46 |
| Dose 2, age in days | 101 | 99,103 | 101 | 99,104 | 100 | 99,103 | 100 | 98,103 | 100 | 98,102 | 100 | 98,103 | 72 | 70,74 |
| Dose 3, age in days | 277 | 273,290 | 276 | 273,284 | 276 | 273,286 | 277 | 274,286 | 276 | 273,289 | 277 | 273,290 | 101 | 99,104 |

**Table S2C: Characteristics of the post-boost per-protocol cohort (n, %)**

| | PCV13 2p+1 | | | | | | PCV10 2p+1 | | | | | |
|---|---|---|---|---|---|---|---|---|---|---|---|---|
| | Full dose | | 40% dose | | 20% dose | | Full dose | | 40% dose | | 20% dose | |
| | n | % | n | % | n | % | n | % | n | % | n | % |
| Season | | | | | | | | | | | | |
| Jan-Mar | 29 | 15 | 26 | 14 | 28 | 15 | 25 | 14 | 28 | 15 | 32 | 17 |
| Apr-Jun | 60 | 32 | 62 | 33 | 63 | 33 | 64 | 35 | 60 | 32 | 61 | 32 |
| Jul-Sep | 49 | 26 | 45 | 24 | 44 | 23 | 46 | 25 | 50 | 27 | 49 | 25 |
| Oct-Dec | 52 | 27 | 55 | 29 | 54 | 29 | 48 | 26 | 50 | 27 | 51 | 26 |
| Year | | | | | | | | | | | | |
| 2019 | 11 | 6 | 10 | 5 | 8 | 4 | 10 | 5 | 8 | 4 | 10 | 5 |
| 2020 | 7 | 4 | 7 | 4 | 11 | 6 | 6 | 3 | 10 | 5 | 9 | 5 |
| 2021 | 172 | 91 | 171 | 91 | 170 | 90 | 167 | 91 | 170 | 90 | 174 | 90 |
| Maternal HIV | | | | | | | | | | | | |
| Positive | 5 | 3 | 6 | 3 | 12 | 6 | 4 | 2 | 5 | 3 | 10 | 5 |
| Negative | 185 | 97 | 182 | 97 | 176 | 93 | 179 | 98 | 182 | 97 | 183 | 95 |
| Unknown | 0 | 0 | 0 | 0 | 1 | 1 | 0 | 0 | 1 | 1 | 0 | 0 |
| Breastfeeding, M10 | | | | | | | | | | | | |
| No | 1 | 1 | 2 | 1 | 4 | 2 | 1 | 1 | 0 | 0 | 1 | 1 |
| Yes | 188 | 99 | 186 | 99 | 185 | 98 | 181 | 99 | 188 | 100 | 192 | 99 |
| Unknown | 1 | 1 | 0 | 0 | 0 | 0 | 1 | 1 | 0 | 0 | 0 | 0 |

| | PCV13 2p+1 | | | | | | PCV10 2p+1 | | | | | |
|---|---|---|---|---|---|---|---|---|---|---|---|---|
| | Full dose | | 40% dose | | 20% dose | | Full dose | | 40% dose | | 20% dose | |
| | n | % | n | % | n | % | n | % | n | % | n | % |
| Sex | | | | | | | | | | | | |
| Male | 93 | 49 | 87 | 46 | 97 | 51 | 93 | 51 | 92 | 49 | 93 | 48 |
| Female | 97 | 51 | 101 | 54 | 92 | 49 | 90 | 49 | 96 | 51 | 100 | 52 |
| Infant HIV | | | | | | | | | | | | |
| Positive | 0 | 0 | 0 | 0 | 0 | 0 | 0 | 0 | 0 | 0 | 0 | 0 |
| Negative | 185 | 97 | 182 | 97 | 176 | 93 | 179 | 98 | 183 | 97 | 183 | 95 |
| Unknown | 5 | 3 | 6 | 3 | 13 | 7 | 4 | 2 | 5 | 3 | 10 | 5 |

Note: Unless otherwise stated characteristics are those recorded at enrolment

**Table S2D: Characteristics of the post-boost per-protocol cohort (median, inter-quartile range)**

| | PCV13 2p+1 | | | | | | PCV10 2p+1 | | | | | |
|---|---|---|---|---|---|---|---|---|---|---|---|---|
| | Full dose | | 40% dose | | 20% dose | | Full dose | | 40% dose | | 20% dose | |
| | Med. | IQR | Med. | IQR | Med. | IQR | Med. | IQR | Med. | IQR | Med. | IQR |
| Maternal age, yrs | 24 | 21,30 | 26 | 21,32 | 26 | 21,32 | 25 | 21,30 | 26 | 21,31 | 26 | 22,30 |
| Infant weight, kgs | 4 | 4,5 | 4 | 4,5 | 5 | 4,5 | 4 | 4,5 | 4 | 4,5 | 4 | 4,5 |
| Dose 1, age in days | 43 | 42,45 | 43 | 42,46 | 43 | 42,46 | 43 | 42,45 | 43 | 42,45 | 43 | 42,45 |
| Dose 2, age in days | 100 | 99,103 | 100 | 99,104 | 100 | 98,103 | 100 | 98,102 | 100 | 98,102 | 100 | 98,102 |
| Dose 3, age in days | 277 | 273,283 | 275 | 272,281 | 275 | 273,283 | 276 | 273,282 | 276 | 273,282 | 276 | 273,283 |

**Table S3A: Percentage of vaccine responders (IgG concentration >=0.35 mcg/ml) post-primary series (18 weeks of age)**

| | PCV13 2p+1 | | | | | | PCV10 2p+1[1] | | | | | | PCV10 3+0[2] | |
|---|---|---|---|---|---|---|---|---|---|---|---|---|---|---|
| | Full dose | | 40% dose | | 20% dose | | Full dose | | 40% dose | | 20% dose | | Full dose | |
| Serotype | n/N | % | n/N | % | n/N | % | n/N | % | n/N | % | n/N | % | n/N | % |
| 1 | 232/232 | 100 | 220/221 | 100 | 221/222 | 100 | 214/218 | 98 | 216/221 | 98 | 212/224 | 95 | 0/0 | |
| 4 | 230/233 | 99 | 221/221 | 100 | 217/222 | 98 | 212/218 | 97 | 213/221 | 96 | 211/224 | 94 | 222/230 | 97 |
| 5 | 225/232 | 97 | 214/221 | 97 | 208/222 | 94 | 203/218 | 93 | 189/220 | 86 | 185/224 | 83 | 0/0 | |
| 6B | 215/232 | 93 | 194/218 | 89 | 170/220 | 77 | 190/218 | 87 | 183/221 | 83 | 186/225 | 83 | 203/230 | 88 |
| 7F | 232/232 | 100 | 220/221 | 100 | 221/222 | 100 | 216/218 | 99 | 214/221 | 97 | 218/224 | 97 | 0/0 | |
| 9V | 226/233 | 97 | 215/221 | 97 | 213/223 | 96 | 211/219 | 96 | 210/221 | 95 | 205/225 | 91 | 223/230 | 97 |
| 14 | 223/232 | 96 | 206/221 | 93 | 199/222 | 90 | 211/218 | 97 | 213/221 | 96 | 215/225 | 96 | 225/229 | 98 |
| 18C | 220/232 | 95 | 210/221 | 95 | 198/222 | 89 | 211/219 | 96 | 212/221 | 96 | 209/224 | 93 | 228/230 | 99 |
| 19F | 231/231 | 100 | 219/219 | 100 | 221/221 | 100 | 215/217 | 99 | 212/213 | 100 | 217/222 | 98 | 226/230 | 98 |
| 23F | 202/230 | 88 | 188/220 | 85 | 169/221 | 76 | 169/218 | 78 | 148/221 | 67 | 151/224 | 67 | 196/230 | 85 |
| 3 | 210/227 | 93 | 188/216 | 87 | 136/219 | 62 | 0/0 | | 0/0 | | 0/0 | | 0/0 | |
| 6A | 208/233 | 89 | 200/220 | 91 | 190/223 | 85 | 0/0 | | 0/0 | | 0/0 | | 0/0 | |
| 19A | 226/231 | 98 | 213/221 | 96 | 214/222 | 96 | 0/0 | | 0/0 | | 0/0 | | 0/0 | |

**Table S3B: Comparison of vaccine responders between study arms post-primary series (18 weeks of age)**

| | PCV13 | | | | PCV10 | | | | | |
|---|---|---|---|---|---|---|---|---|---|---|
| | 40% dose - Full dose | | 20% dose - Full dose | | 40% dose - Full dose[1] | | 20% dose - Full dose[1] | | 2p+1 - 3p+0[2] | |
| Serotype | RD | 90% CI | RD | 90% CI | RD | 90% CI | RD | 90% CI | RD | 90% CI |
| 1 | -0.5 | -1.7, 0.8 | -0.5 | -1.7, 0.8 | -0.4 | -2.8, 2.0 | -3.5 | -6.5, -0.4 | | |
| 4 | 1.3 | -0.3, 2.8 | -1.0 | -3.2, 1.3 | -0.9 | -3.8, 2.1 | -3.1 | -6.3, 0.3 | 0.7 | -2.2, 3.6 |
| 5 | -0.2 | -3.0, 2.7 | -3.3 | -6.6, 0.1 | -7.2 | -12.0, -2.3 | -10.5 | -15.5, -5.4 | | |
| 6B | -3.7 | -8.2, 0.9 | -15.4 | -20.7, -9.8 | -4.4 | -9.9, 1.3 | -4.5 | -10.0, 1.2 | -1.1 | -6.3, 4.0 |
| 7F | -0.5 | -1.7, 0.8 | -0.5 | -1.7, 0.8 | -2.2 | -4.6, 0.2 | -1.8 | -4.0, 0.6 | | |
| 9V | 0.3 | -2.5, 3.0 | -1.5 | -4.5, 1.6 | -1.3 | -4.6, 2.0 | -5.2 | -9.0, -1.3 | -0.6 | -3.6, 2.3 |
| 14 | -2.9 | -6.5, 0.7 | -6.5 | -10.5, -2.4 | -0.4 | -3.4, 2.6 | -1.2 | -4.3, 1.9 | -1.5 | -4.1, 1.1 |
| 18C | 0.2 | -3.3, 3.7 | -5.6 | -9.9,-1.4 | -0.4 | -3.6, 2.7 | -3.0 | -6.6, 0.6 | -2.8 | -5.3, -0.3 |
| 19F | 0.0 | -1.0, 1.0 | 0.0 | -1.0, 1.0 | 0.5 | -1.2, 2.1 | -1.3 | -3.5, 0.9 | 0.8 | -1.2, 2.8 |
| 23F | -2.4 | -7.7, 2.9 | -11.4 | -17.2, -5.4 | -10.6 | -17.4, -3.5 | -10.1 | -16.9, -3.1 | -7.7 | -13.7, -1.6 |
| 3 | -5.5 | -10.2, -0.7 | -30.4 | -36.3,-24.0 | | | | | | |
| 6A | 1.6 | -3.1, 6.3 | -4.1 | -9.2, 1.1 | | | | | | |
| 19A | -1.5 | -4.2, 1.3 | -1.4 | -4.2, 1.3 | | | | | | |

Table 2A, 2B notes: [1] Samples from participants in the PCV10 2p+1 arms were assayed for IgG to the 10 PCV10 serotypes.
[2] Samples from the participants in the PCV10 3p+0 arm were assayed for IgG to 7 (PCV7) serotypes due to funding constraints for this secondary endpoint.

**Table S4A: Geometric mean IgG concentrations post-boost (approx. 10 months of age)**

| | PCV13 2p+1 | | | | | | PCV10 2p+1 | | | | | |
|---|---|---|---|---|---|---|---|---|---|---|---|---|
| | Full dose | | 40% dose | | 20% dose | | Full dose | | 40% dose | | 20% dose | |
| **Serotype** | **N** | **GMC** | **N** | **GMC** | **N** | **GMC** | **N** | **GMC** | **N** | **GMC** | **N** | **GMC** |
| 1 | 190 | 4.4 | 188 | 3.4 | 189 | 2.6 | 180 | 3.1 | 180 | 1.7 | 182 | 1.3 |
| 4 | 190 | 3.3 | 188 | 2.3 | 189 | 1.9 | 183 | 2.8 | 188 | 2.0 | 191 | 1.5 |
| 5 | 190 | 1.9 | 188 | 1.5 | 189 | 1.3 | 183 | 0.8 | 187 | 0.5 | 192 | 0.4 |
| 6B | 190 | 12.9 | 188 | 11.2 | 189 | 7.5 | 183 | 4.5 | 188 | 3.0 | 193 | 2.3 |
| 7F | 190 | 7.2 | 188 | 5.3 | 189 | 4.3 | 183 | 3.5 | 188 | 2.3 | 193 | 1.7 |
| 9V | 189 | 4.0 | 188 | 3.0 | 187 | 2.7 | 181 | 2.8 | 186 | 1.9 | 188 | 1.4 |
| 14 | 190 | 7.6 | 188 | 6.3 | 189 | 4.3 | 183 | 4.5 | 188 | 2.7 | 193 | 1.8 |
| 18C | 190 | 3.6 | 188 | 2.5 | 189 | 1.8 | 183 | 7.8 | 188 | 4.5 | 192 | 3.2 |
| 19F | 190 | 13.1 | 186 | 10.6 | 188 | 9.8 | 180 | 14.8 | 184 | 12.2 | 188 | 9.5 |
| 23F | 189 | 4.9 | 186 | 4.0 | 187 | 3.0 | 174 | 1.7 | 185 | 1.0 | 186 | 0.7 |
| 3 | 186 | 1.0 | 187 | 0.8 | 189 | 0.7 | 0 | | 0 | | 0 | |
| 6A | 190 | 9.9 | 188 | 7.0 | 189 | 5.7 | 0 | | 0 | | 0 | |
| 19A | 187 | 9.3 | 184 | 7.5 | 188 | 6.1 | 0 | | 0 | | 0 | |

**Table S4B: Comparison of geometric mean antibody concentrations post-boost between study arms (approx. 10 months of age)**

| | PCV13 | | | | PCV10 | | | |
|---|---|---|---|---|---|---|---|---|
| | 40% dose / Full dose | | 20% dose / Full dose | | 40% dose / Full dose | | 20% dose / Full dose | |
| **Serotype** | **Ratio** | **90% CI** | **Ratio** | **90% CI** | **Ratio** | **90% CI** | **Ratio** | **90% CI** |
| 1 | 0.76 | 0.67,0.87 | 0.59 | 0.52,0.67 | 0.57 | 0.48,0.66 | 0.42 | 0.36,0.50 |
| 4 | 0.72 | 0.63,0.82 | 0.58 | 0.51,0.65 | 0.70 | 0.62,0.79 | 0.53 | 0.47,0.61 |
| 5 | 0.77 | 0.68,0.87 | 0.66 | 0.58,0.74 | 0.69 | 0.62,0.78 | 0.54 | 0.48,0.60 |
| 6B | 0.87 | 0.76,0.99 | 0.58 | 0.51,0.68 | 0.65 | 0.58,0.74 | 0.51 | 0.46,0.58 |
| 7F | 0.73 | 0.66,0.82 | 0.60 | 0.54,0.66 | 0.66 | 0.59,0.75 | 0.48 | 0.42,0.54 |
| 9V | 0.76 | 0.67,0.87 | 0.68 | 0.60,0.77 | 0.66 | 0.59,0.74 | 0.49 | 0.43,0.55 |
| 14 | 0.83 | 0.69,1.00 | 0.57 | 0.47,0.69 | 0.59 | 0.48,0.72 | 0.40 | 0.33,0.50 |
| 18C | 0.69 | 0.60,0.79 | 0.50 | 0.44,0.57 | 0.58 | 0.50,0.67 | 0.42 | 0.36,0.48 |
| 19F | 0.81 | 0.70,0.93 | 0.75 | 0.64,0.87 | 0.83 | 0.70,0.97 | 0.64 | 0.55,0.76 |
| 23F | 0.80 | 0.68,0.95 | 0.61 | 0.51,0.72 | 0.58 | 0.50,0.68 | 0.41 | 0.35,0.48 |
| 3 | 0.84 | 0.74,0.97 | 0.76 | 0.66,0.88 | | | | |
| 6A | 0.71 | 0.61,0.82 | 0.57 | 0.49,0.67 | | | | |
| 19A | 0.81 | 0.69,0.94 | 0.66 | 0.56,0.78 | | | | |

**Table S4C: Geometric mean titres of opsonophagocytic activity (OPA) post-boost (approx. 10 months of age)**

| | PCV13 2p+1 | | | | | | PCV10 2p+1 | | | | | |
|---|---|---|---|---|---|---|---|---|---|---|---|---|
| | Full dose | | 40% dose | | 20% dose | | Full dose | | 40% dose | | 20% dose | |
| **Serotype** | **N** | **GMT** | **N** | **GMT** | **N** | **GMT** | **N** | **GMT** | **N** | **GMT** | **N** | **GMT** |
| 1 | 48 | 266 | 50 | 217 | 49 | 204 | 45 | 271 | 49 | 147 | 48 | 120 |
| 4 | 49 | 1,821 | 48 | 1,640 | 50 | 1,784 | 45 | 1,361 | 47 | 857 | 47 | 602 |
| 5 | 49 | 496 | 50 | 387 | 50 | 392 | 48 | 461 | 48 | 313 | 49 | 213 |
| 6B | 49 | 2,749 | 50 | 2,898 | 50 | 2,103 | 48 | 1,007 | 47 | 610 | 49 | 481 |
| 7F | 49 | 4,112 | 50 | 2,788 | 50 | 2,938 | 47 | 2,183 | 49 | 1,482 | 49 | 1,153 |
| 9V | 49 | 1,448 | 50 | 1,105 | 50 | 735 | 48 | 656 | 49 | 338 | 49 | 262 |
| 14 | 49 | 1,383 | 49 | 859 | 50 | 1,359 | 46 | 1,069 | 48 | 430 | 49 | 270 |
| 18C | 49 | 1,325 | 50 | 1,131 | 50 | 872 | 48 | 3,584 | 49 | 2,164 | 49 | 1,777 |
| 19F | 49 | 1,037 | 50 | 842 | 50 | 732 | 47 | 1,182 | 49 | 972 | 49 | 446 |
| 23F | 49 | 3,047 | 50 | 3,706 | 50 | 3,541 | 48 | 797 | 49 | 340 | 46 | 336 |

**Table S4D: Percentage of OPA titres >8 post-boost (approx. 10 months of age)**

| | PCV13 2p+1 | | | | | | PCV10 2p+1 | | | | | |
|---|---|---|---|---|---|---|---|---|---|---|---|---|
| | Full dose | | 40% dose | | 20% dose | | Full dose | | 40% dose | | 20% dose | |
| **Serotype** | **n/N** | **%** | **n/N** | **%** | **n/N** | **%** | **n/N** | **%** | **n/N** | **%** | **n/N** | **%** |
| 1 | 48/48 | 100 | 49/50 | 98 | 47/49 | 96 | 44/45 | 98 | 46/49 | 94 | 44/48 | 92 |
| 4 | 48/49 | 98 | 48/48 | 100 | 50/50 | 100 | 45/45 | 100 | 47/47 | 100 | 46/47 | 98 |
| 5 | 49/49 | 100 | 50/50 | 100 | 50/50 | 100 | 47/48 | 98 | 47/48 | 98 | 48/49 | 98 |
| 6B | 49/49 | 100 | 50/50 | 100 | 49/50 | 98 | 48/48 | 100 | 47/47 | 100 | 48/49 | 98 |
| 7F | 49/49 | 100 | 50/50 | 100 | 50/50 | 100 | 47/47 | 100 | 49/49 | 100 | 49/49 | 100 |
| 9V | 49/49 | 100 | 50/50 | 100 | 50/50 | 100 | 48/48 | 100 | 49/49 | 100 | 48/49 | 98 |
| 14 | 47/49 | 96 | 44/49 | 90 | 49/50 | 98 | 46/46 | 100 | 42/48 | 88 | 41/49 | 84 |
| 18C | 49/49 | 100 | 50/50 | 100 | 50/50 | 100 | 48/48 | 100 | 49/49 | 100 | 49/49 | 100 |
| 19F | 49/49 | 100 | 49/50 | 98 | 48/50 | 96 | 44/47 | 94 | 48/49 | 98 | 43/49 | 88 |
| 23F | 49/49 | 100 | 50/50 | 100 | 50/50 | 100 | 48/48 | 100 | 49/49 | 100 | 46/46 | 100 |

**Table S4E: Comparison of responders (OPA titre >8) between study arms post-boost (approx. 10 months of age)**

| | PCV13 | | | | PCV10 | | | |
|---|---|---|---|---|---|---|---|---|
| | 40% dose - Full dose | | 20% dose - Full dose | | 40% dose - Full dose | | 20% dose - Full dose | |
| **Serotype** | **RD** | **90% CI** | **RD** | **90% CI** | **RD** | **90% CI** | **RD** | **90% CI** |
| 1 | -2.0 | -7.3, 3.6 | -4.1 | -10.2, 2.4 | -3.9 | -11.4, 4.3 | -6.1 | -14.2, 2.7 |
| 4 | 2.0 | -3.6, 7.5 | 2.0 | -3.5, 7.5 | 0.0 | -4.7, 4.9 | -2.1 | -7.8, 3.8 |
| 5 | 0.0 | -4.4, 4.5 | 0.0 | -4.4, 4.5 | 0.0 | -6.4, 6.4 | 0.0 | -6.3, 6.5 |
| 6B | 0.0 | -4.4, 4.5 | -2.0 | -7.3, 3.5 | 0.0 | -4.7, 4.6 | -2.0 | -7.5, 3.6 |
| 7F | 0.0 | -4.4, 4.5 | 0.0 | -4.4, 4.5 | 0.0 | -4.5, 4.7 | 0.0 | -4.5, 4.7 |
| 9V | 0.0 | -4.4, 4.5 | 0.0 | -4.4, 4.5 | 0.0 | -4.5, 4.6 | -2.0 | -7.5, 3.6 |
| 14 | -6.1 | -15.1, 3.3 | 2.1 | -4.9, 9.0 | -12.5 | -20.7, -3.2 | -16.3 | -25.0, -6.2 |
| 18C | 0.0 | -4.4, 4.5 | 0.0 | -4.4, 4.5 | 0.0 | -4.5, 4.6 | 0.0 | -4.5, 4.6 |
| 19F | -2.0 | -7.3, 3.5 | -4.0 | -10.0, 2.4 | 4.3 | -3.6,12.1 | -5.9 | -15.8, 4.6 |
| 23F | 0.0 | -4.4, 4.5 | 0.0 | -4.4, 4.5 | 0.0 | -4.5, 4.6 | 0.0 | -4.8, 4.6 |

**Table S5A: Serotype-specific carriage prevalence at 9 months of age**

| | PCV13 2p+1 | | | | | | PCV10 2p+1 | | | | | | PCV10 3+0 | |
|---|---|---|---|---|---|---|---|---|---|---|---|---|---|---|
| | Full dose | | 40% dose | | 20% dose | | Full dose | | 40% dose | | 20% dose | | Full dose | |
| | N=207 | | N=210 | | N=206 | | N=198 | | N=203 | | N=209 | | N=206 | |
| **Serotype** | **n** | **%** | **n** | **%** | **n** | **%** | **n** | **%** | **n** | **%** | **n** | **%** | **n** | **%** |
| 1 | 0 | 0.0 | 0 | 0.0 | 0 | 0.0 | 0 | 0.0 | 0 | 0.0 | 0 | 0.0 | 0 | 0.0 |
| 4 | 0 | 0.0 | 1 | 0.5 | 0 | 0.0 | 0 | 0.0 | 0 | 0.0 | 0 | 0.0 | 0 | 0.0 |
| 5 | 0 | 0.0 | 0 | 0.0 | 0 | 0.0 | 0 | 0.0 | 0 | 0.0 | 0 | 0.0 | 0 | 0.0 |
| 6B | 0 | 0.0 | 0 | 0.0 | 0 | 0.0 | 0 | 0.0 | 0 | 0.0 | 1 | 0.5 | 0 | 0.0 |
| 7F | 0 | 0.0 | 0 | 0.0 | 0 | 0.0 | 0 | 0.0 | 0 | 0.0 | 0 | 0.0 | 0 | 0.0 |
| 9V | 0 | 0.0 | 0 | 0.0 | 1 | 0.5 | 0 | 0.0 | 0 | 0.0 | 0 | 0.0 | 0 | 0.0 |
| 14 | 0 | 0.0 | 3 | 1.4 | 3 | 1.5 | 3 | 1.5 | 3 | 1.5 | 2 | 1.0 | 4 | 1.9 |
| 18C | 0 | 0.0 | 0 | 0.0 | 0 | 0.0 | 0 | 0.0 | 0 | 0.0 | 0 | 0.0 | 0 | 0.0 |
| 19F | 10 | 4.8 | 10 | 4.8 | 8 | 3.9 | 7 | 3.5 | 8 | 3.9 | 11 | 5.3 | 3 | 1.5 |
| 23F | 0 | 0.0 | 2 | 1.0 | 1 | 0.5 | 0 | 0.0 | 1 | 0.5 | 4 | 1.9 | 2 | 1.0 |
| 3 | 7 | 3.4 | 5 | 2.4 | 5 | 2.4 | 6 | 3.0 | 11 | 5.4 | 5 | 2.4 | 8 | 3.9 |
| 6A | 10 | 4.8 | 17 | 8.1 | 12 | 5.8 | 16 | 8.1 | 13 | 6.4 | 22 | 10.5 | 19 | 9.2 |
| 19A | 10 | 4.8 | 11 | 5.2 | 13 | 6.3 | 17 | 8.6 | 13 | 6.4 | 16 | 7.7 | 16 | 7.8 |
| PCV13 | 37 | 17.9 | 49 | 23.3 | 43 | 20.9 | 49 | 24.7 | 49 | 24.1 | 61 | 29.2 | 52 | 25.2 |
| PCV10 | 10 | 4.8 | 16 | 7.6 | 13 | 6.3 | 10 | 5.1 | 12 | 5.9 | 18 | 8.6 | 9 | 4.4 |
| 3/6A/19A | 27 | 13.0 | 33 | 15.7 | 30 | 14.6 | 39 | 19.7 | 37 | 18.2 | 43 | 20.6 | 43 | 20.9 |
| 6A/19A | 20 | 9.7 | 28 | 13.3 | 25 | 12.1 | 33 | 16.7 | 26 | 12.8 | 38 | 18.2 | 35 | 17.0 |
| Any carriage | 174 | 84.1 | 173 | 82.4 | 168 | 81.6 | 158 | 79.8 | 167 | 82.3 | 178 | 85.2 | 173 | 84.0 |

### Table S5B: Serotype-specific carriage prevalence at 18 months of age

| | PCV13 2p+1 | | | | | | PCV10 2p+1 | | | | | | PCV10 3+0 | |
|---|---|---|---|---|---|---|---|---|---|---|---|---|---|---|
| | Full dose | | 40% dose | | 20% dose | | Full dose | | 40% dose | | 20% dose | | Full dose | |
| | N=193 | | N=196 | | N=191 | | N=179 | | N=194 | | N=190 | | N=221 | |
| **Serotype** | **n** | **%** | **n** | **%** | **n** | **%** | **n** | **%** | **n** | **%** | **n** | **%** | **n** | **%** |
| 1 | 0 | 0.0 | 0 | 0.0 | 0 | 0.0 | 0 | 0.0 | 0 | 0.0 | 1 | 0.5 | 0 | 0.0 |
| 4 | 0 | 0.0 | 1 | 0.5 | 0 | 0.0 | 0 | 0.0 | 0 | 0.0 | 0 | 0.0 | 0 | 0.0 |
| 5 | 0 | 0.0 | 0 | 0.0 | 0 | 0.0 | 0 | 0.0 | 0 | 0.0 | 0 | 0.0 | 0 | 0.0 |
| 6B | 0 | 0.0 | 0 | 0.0 | 0 | 0.0 | 0 | 0.0 | 0 | 0.0 | 1 | 0.5 | 0 | 0.0 |
| 7F | 0 | 0.0 | 0 | 0.0 | 0 | 0.0 | 0 | 0.0 | 0 | 0.0 | 0 | 0.0 | 0 | 0.0 |
| 9V | 2 | 1.0 | 0 | 0.0 | 0 | 0.0 | 0 | 0.0 | 0 | 0.0 | 1 | 0.5 | 0 | 0.0 |
| 14[1] | 1 | 0.5 | 0 | 0.0 | 2 | 1.0 | 1 | 0.6 | 4 | 2.1 | 10 | 5.3 | 6 | 2.7 |
| 18C | 1 | 0.5 | 0 | 0.0 | 0 | 0.0 | 0 | 0.0 | 0 | 0.0 | 0 | 0.0 | 1 | 0.5 |
| 19F[2] | 6 | 3.1 | 14 | 7.1 | 11 | 5.8 | 5 | 2.8 | 6 | 3.1 | 7 | 3.7 | 14 | 6.3 |
| 23F | 0 | 0.0 | 2 | 1.0 | 0 | 0.0 | 0 | 0.0 | 1 | 0.5 | 0 | 0.0 | 0 | 0.0 |
| 3 | 5 | 2.6 | 7 | 3.6 | 8 | 4.2 | 6 | 3.4 | 10 | 5.2 | 9 | 4.7 | 11 | 5.0 |
| 6A | 7 | 3.6 | 8 | 4.1 | 2 | 1.0 | 11 | 6.1 | 19 | 9.8 | 17 | 8.9 | 12 | 5.4 |
| 19A[3] | 12 | 6.2 | 5 | 2.6 | 8 | 4.2 | 11 | 6.1 | 13 | 6.7 | 6 | 3.2 | 8 | 3.6 |
| PCV13 | 34 | 17.6 | 37 | 18.9 | 31 | 16.2 | 34 | 19.0 | 53 | 27.3 | 52 | 27.4 | 52 | 23.5 |
| PCV10[4] | 10 | 5.2 | 17 | 8.7 | 13 | 6.8 | 6 | 3.4 | 11 | 5.7 | 20 | 10.5 | 21 | 9.5 |
| 3/6A/19A | 24 | 12.4 | 20 | 10.2 | 18 | 9.4 | 28 | 15.6 | 42 | 21.6 | 32 | 16.8 | 31 | 14.0 |
| 6A/19A | 19 | 9.8 | 13 | 6.6 | 10 | 5.2 | 22 | 12.3 | 32 | 16.5 | 23 | 12.1 | 20 | 9.0 |
| Any carriage | 149 | 77.2 | 149 | 76.0 | 129 | 67.5 | 133 | 74.3 | 140 | 72.2 | 137 | 72.1 | 160 | 72.4 |

**Table S5C: Differences between study arms in carriage at 9 months of age**

| Comparison | Serotypes | RD | 90% CI | p-value[1] |
|---|---|---|---|---|
| PCV13 20% dose vs full | PCV13 | 5.5 | -1.1,11.9 | 0.29 |
| PCV13 40% dose vs full | PCV13 | 3.0 | -3.4, 9.4 | 0.54 |
| PCV10 20% dose vs full | PCV10 | 0.9 | -3.0, 4.7 | 0.83 |
| PCV10 40% dose vs full | PCV10 | 3.6 | -0.7, 7.7 | 0.24 |
| PCV13 vs PCV10 (both full dose) | 3/6A/19A | -6.7 | -12.7, -0.5 | 0.14 |
| PCV13 vs PCV10 (both full dose) | 6A/19A | -7.0 | -12.5, -1.4 | 0.08 |
| PCV10 2p+1 vs 3+0 (both full dose) | PCV10 | 0.7 | -2.9, 4.3 | 0.82 |

[1]Fishers exact test

**Table S5D: Differences between study arms in carriage at 18 months of age**

| Comparison | Serotypes | RD | 90% CI | p-value[1] |
|---|---|---|---|---|
| PCV13 20% dose vs full | PCV13 | -1.4 | -4.2, 1.4 | 0.79 |
| PCV13 40% dose vs full | PCV13 | 1.3 | -1.7, 4.3 | 0.79 |
| PCV10 20% dose vs full | PCV10 | 7.1 | 5.9, 8.3 | 0.008 |
| PCV10 40% dose vs full | PCV10 | 2.3 | 1.6, 3.0 | 0.33 |
| PCV13 vs PCV10 (both full dose) | 3/6A/19A | -3.2 | -5.5, -0.9 | 0.46 |
| PCV13 vs PCV10 (both full dose) | 6A/19A | -2.5 | -4.3, -0.7 | 0.51 |
| PCV10 2p+1 vs 3+0 (both full dose) | PCV10 | -6.1 | -7.2, -5.1 | 0.016 |

[1]Fishers exact test

**Table S6. Serious Adverse Events (SAEs), Severe pneumonia SAEs, and adverse events, by study arm**

| | PCV13 2p+1 | | | | | | PCV10 2p+1 | | | | | | PCV10 3+0 | | |
|---|---|---|---|---|---|---|---|---|---|---|---|---|---|---|---|
| | Full dose | | 40% dose | | 20% dose | | Full dose | | 40% dose | | 20% dose | | Full dose | | |
| | n | % | n | % | n | % | n | % | n | % | n | % | n | % | p-value |
| **SAE[1]** | | | | | | | | | | | | | | | 0.69 |
| None | 293 | 98 | 291 | 97 | 285 | 95 | 293 | 98 | 293 | 98 | 291 | 97 | 292 | 97 | |
| 1 | 6 | 2 | 8 | 3 | 12 | 4 | 6 | 2 | 6 | 2 | 9 | 3 | 7 | 2 | |
| 2 | 1 | 0 | 0 | 0 | 2 | 1 | 1 | 0 | 0 | 0 | 0 | 0 | 0 | 0 | |
| 3 | 0 | 0 | 0 | 0 | 0 | 0 | 0 | 0 | 0 | 0 | 0 | 0 | 1 | 0 | |
| Severe pneumonia SAE | 3 | 1 | 5 | 2 | 7 | 2 | 6 | 2 | 4 | 1 | 6 | 2 | 9 | 3 | |
| Severe pneumonia SAE with abnormal CXR | 1 | 0 | 3 | 1 | 1 | 0 | 1 | 0 | 1 | 0 | 1 | 0 | 5 | 2 | |
| Died | | | | | | | | | | | | | | | 0.84 |
| No | 299 | 100 | 298 | 100 | 298 | 100 | 298 | 99 | 299 | 100 | 298 | 99 | 300 | 100 | |
| Yes | 1 | 0 | 1 | 0 | 1 | 0 | 2 | 1 | 0 | 0 | 2 | 1 | 0 | 0 | |
| **All Respiratory AE[2]** | | | | | | | | | | | | | | | 0.70 |
| 0 | 165 | 55 | 148 | 49 | 151 | 51 | 143 | 48 | 160 | 54 | 145 | 48 | 160 | 53 | |
| 1-3 | 109 | 36 | 123 | 41 | 123 | 41 | 134 | 45 | 117 | 39 | 130 | 43 | 110 | 37 | |
| 4-11 | 26 | 9 | 28 | 9 | 25 | 8 | 23 | 8 | 22 | 7 | 25 | 8 | 30 | 10 | |
| Pneumonia AEs | 8 | 3 | 16 | 5 | 8 | 3 | 9 | 3 | 5 | 2 | 7 | 2 | 8 | 3 | |
| Pneumonia AEs with abnormal CXR | 1 | 0 | 4 | 1 | 4 | 1 | 2 | 1 | 1 | 0 | 0 | 0 | 2 | 1 | |

Abbreviations: AE: adverse event; CXR: chest x-ray; SAE: serious adverse event.

Table notes:

[1] Safety reports include ‘initial reports’ of events only. SAEs reported here are all SAEs. Severe pneumonia SAEs are a subcategory of all SAEs, severe pneumonia SAEs with abnormal chest x-ray are a further sub-category of severe pneumonia SAEs. Deaths were reported as SAEs. Severe pneumonia was defined using the WHO Pocket Book for Hospital care of Children (2013) i.e. cough or difficulty breathing/ fast breathing and some evidence of lower chest wall indrawing/ nasal flaring/ grunting (with or without other danger signs). Fast breathing was defined as $\geq$50 breaths per minute in 2-12 months olds and $\geq$40 breaths per minute is >12 months of age.

[2] In March 2020 a protocol amendment was approved to restrict reporting of AEs to only those affecting the respiratory tract to attempt to manage workload more effectively. Most of these were very minor coughs and colds, Pneumonia AEs were cases of non-severe pneumonia and are a subcategory of all AEs and underwent investigation as per SOPs with chest x-ray and a nasopharyngeal swab. Non-severe pneumonia cases were cases of cough or difficulty breathing without any other danger sign.
